# Optimal thromboprophylaxis strategies in non-critically ill patients with COVID-19 pneumonia. The PROTHROMCOVID Randomized Controlled Trial

**DOI:** 10.1101/2022.05.03.22274594

**Authors:** Nuria Muñoz-Rivas, Jesús Aibar, Cristina Gabara-Xancó, Ángela Trueba-Vicente, Ana Urbelz-Pérez, Vicente Gómez-Del Olmo, Pablo Demelo-Rodriguez, Alberto Rivera-Gallego, Pau Bosch-Nicolau, Montserrat Perez-Pinar, Mónica Rios-Prego, Olga Madridano-Cobo, Laura Ramos-Alonso, Jesús Alonso-Carrillo, Iria Francisco-Albelsa, Edelmira Martí-Saez, Ana Maestre-Peiró, Manuel Méndez-Bailón, José Ángel Hernández-Rivas, Juan Torres-Macho, The PROTHROMCOVID Trial investigators

## Abstract

**Background:** Hospitalized patients with COVID-19 are at increased risk for thrombosis, acute respiratory distress syndrome and death. The optimal dosage of thromboprophylaxis is unknown.

**Objective:** To evaluate the efficacy and safety of tinzaparin in prophylactic, intermediate, and therapeutic doses in non-critical patients admitted for COVID-19 pneumonia.

**Design, setting, and participants:** Randomized controlled, multicenter trial (PROTHROMCOVID) enrolling non-critical, hospitalized adult patients with COVID-19 pneumonia.

**Interventions:** Patients were randomized to prophylactic (4500 IU), intermediate (100 IU/kg), or therapeutic (175 IU/kg) doses of tinzaparin during hospitalization, followed by 7 days of prophylactic tinzaparin at discharge.

**Measurements:** The primary efficacy outcome was a composite endpoint of symptomatic systemic thrombotic events, need for invasive or non-invasive mechanical ventilation, or death within 30 days. The main safety outcome was major bleeding at 30 days.

**Results:** Of the 311 subjects randomized, 300 were included in the analysis (mean [SD] age, 56.7 [14.6] years; males, 182 [60.7%]. The composite endpoint at 30 days from randomization occurred in 58 patients (19.3%) of the total population; 19 (17.1 %) in the prophylactic group, 20 (22.1%) in the intermediate group, and 19 (18.5%) in the therapeutic dose group (P= 0.72). No major bleeding event was reported; non-major bleeding was observed in 3.7% of patients, with no intergroup differences.

**Conclusions:** In non-critically ill COVID-19 patients, intermediate or full-dose tinzaparin compared to standard prophylactic doses did not appear to increase benefit regarding the likelihood of thrombotic event, non-invasive ventilation or high-flow oxygen, or death.

**Trial Registration:** ClinicalTrials.gov Identifier (NCT04730856).

**Funding:** This independent research initiative was supported by Leo-Pharma; Tinzaparin was provided by Leo Pharma.

## Introduction

Severe, acute, respiratory syndrome-coronavirus 2 (SARS-CoV-2) infection can cause different clinical manifestations, ranging from mild to very severe symptomatology, with significant morbidity and mortality, principally associated with bilateral pneumonia that can cause acute respiratory distress syndrome (ARSD). More than 6 million people have died since the first reports in late 2019 in Wuhan, China and it is estimated that more than 450 million people have been infected with COVID-19 to date. Recent research estimates more than that 18 million people died worldwide because of the COVID-19 pandemic (as measured by excess mortality) over that period(1). Since the first wave of the SARS-CoV-2 pandemic, the increase in systemic thrombosis in hospitalized patients was evident (2), particularly in critical care units worldwide (3)(4). The phenomenon known as ‘pulmonary immunothrombosis’ correlated with the severity of respiratory failure and need for ventilation in individuals with COVID-19(5). The association of viral infection with thrombosis is mediated by two interrelated processes: a state of hypercoagulability that causes large vessel thrombosis and direct endothelial damage that provokes *in situ* thrombosis. Subsequently, more and more evidence has been published of the so-called ‘COVID-19-associated coagulopathy’. It was then hypothesized that anticoagulation could improve the life expectancy of patients with SARS-CoV-2 infection who, given the severity of their disease, required hospitalization. At the beginning of the pandemic, while awaiting the results of clinical trials, different protocols of prophylactic anticoagulation have been developed in hospitals. These included the use of standard, intermediate and even full doses of low-molecular weight heparin (LMWH) (6). This was the underlying premise for conducting numerous clinical studies to evaluate the efficacy and safety of therapeutic or intermediate doses (with either LMWH or different oral anticoagulants) versus prophylactic doses of anticoagulation. The results of several clinical trials have been published to date, focusing on anticoagulation intensity in patients admitted for COVID-19 (7). While the results have been more robust against higher than usual prophylactic doses (8) in the critically ill patients(9), uncertainty persists as to optimal LMWH doses in non-critical cases. Most trials have evaluated standard prophylactic LMWH dose strategies versus therapeutic doses or other oral anticoagulants, with contradictory results (10)(11).

The PROTHROMCOVID multicenter clinical trial was carried out to evaluate the efficacy of tinzaparin treatment at different doses (prophylactic, intermediate, and therapeutic) in patients with COVID-19 pneumonia to probe the endpoints of death, need for mechanical ventilation and venous or arterial thrombosis within 30 days following randomization. This trial also examined the safety of tinzaparin at different doses in relation to developing complications of both major and minor bleeding.

## Material and methods

### Study Design

The PROTHROMCOVID study (NCT04730856) is a randomized, open-label, multicenter, controlled study in hospitalized patients with COVID-19 pneumonia, conducted in conventional hospital wards in 18 academic hospitals in Spain. This investigator-initiated clinical trial enrolled individuals with COVID-19 pneumonia who were hospitalized from February 1st, 2021 to September 30th, 2021.

### Patients

Adults with a body weight of 50-100 kg who required admission to a conventional (non-critical) hospital ward due to COVID-19 pneumonia were included if they also met any of the following criteria: a) baseline oxygen saturation ≤94%, b) D-dimer > 1000 μg/L,c) C Reactive Protein (CRP) > 150 mg/L, or d) interleukin-6 (IL6) > 40 pg/mL). The main exclusion criteria were: a) need for full-dose anticoagulant therapy, b) active bleeding or situations prone to bleeding, c) glomerular filtration rate < 30 ml/min/1.73 m^2^, d) platelet count<80 × 10^9^/L, e) previous heparin-induced thrombocytopenia, and f) hypersensitivity/intolerance to heparins. The study design and full list of eligibility and exclusion criteria are provided in the supplementary material (Supplementary file Table S1 and Table S2)

### Randomization

Patients were screened at hospitalization and randomized at a ratio of 1:1:1 by means of a central, electronic, automated system with permuted blocks of 6. Neither participants nor investigators were blinded as to group assignment. Subjects were stratified by age, sex, and presence of high blood pressure. Those who were assigned to the control group received standard prophylaxis with subcutaneous (sc) tinzaparin 4500 IU once daily. The experimental group received tinzaparin 100 IU/kg once daily (intermediate dose group) or 175 IU/kg once daily (therapeutic dose group). The assigned treatments remained the same throughout hospitalization. After discharge, all patients received tinzaparin 4500 IU/day subcutaneously for seven days, after which thromboprophylaxis was maintained at the discretion of the investigator. If intensive care unit (ICU) admission was required, the patients could remain with the study drug or not, according to local practices. Except for the assigned anticoagulation therapy, all other clinical care was provided as per local protocols.

### Outcomes

Demographic characteristics, comorbidities, medications, and laboratory evaluations were recorded at randomization. The primary efficacy outcome was a composite endpoint of death, need for invasive mechanical ventilation (IMV), non-invasive ventilatation (NIV) or high flow oxygen with nasal cannula (HFNC), and venous or arterial thrombosis within 30 days after randomization. Secondary efficacy variables were the same endpoints at 30 and 90 days, progression to ARDS, and length of hospital stay. Safety outcomes were major bleeding and clinically relevant non-major bleeding, as defined by the International Society on Thrombosis and Hemostasis (ISTH) (12). Outcomes were adjudicated locally by the investigators based on objectively confirmed diagnostic tests, laboratory results, and other objective data from the clinical record.

### Statistical analysis

Considering the main objective, the incidence in the control group was expected to be 24% and 12% in groups 2 and 3. The sample size was calculated from a proportion of a 13% reduction in thrombosis in the control group and a difference of 5% compared to treatment groups.

Accepting an alpha risk of 0.025 and a beta risk of <0.2 in a bilateral contrast, statistically significant differences could be detected with 200 patients per group. The study protocol included an interim analysis when 50% of the target population had been included. An interim analysis was scheduled to be performed after 300 patients were included. The trial could be stopped for: (1) superiority; (2) futility with regard to the primary endpoint; or (3) safety reasons. Following the results of this interim analysis presented in this article, the Scientific Committee decided to prematurely halt the clinical trial, based on the futility analysis and the drop in recruitment at the end of fifth wave.

Categorical variables were expressed as frequencies and percentages, and quantitative variables as mean ± standard deviation (SD) or median and interquartile range (IQR), relative to distribution. The Saphiro-Wilk test was used to examine the normality of the distributions of samples of <30 and the Kolmogorov-Smirnoff test was applied in the other cases. For intergroup statistical analysis, chi-square or Fisher’s exact test were used for categorical variables and unpaired Student’s t test or Mann-Whitney test for continuous variables. Survival analysis was carried out using Kaplan-Meier curves. Efficacy and safety were assessed in the intention-to-treat population, including all randomized patients who received at least one dose of the assigned treatment. Statistical analyses were performed using the statistical package SAS, 9.4 (Copyright © 2016 by SAS Institute Inc., Cary, NC, USA).

#### Ethical Issues

The Foundation for Biomedical Research and Innovation of the Infanta Leonor University Hospital and the Southeast University Hospital sponsored the study. The trial was conducted in accordance with the Declaration of Helsinki and local regulatory requirements. The protocol was approved by the Ethical Committees for Drug Research at the Gregorio Marañón General University Hospital and, subsequently, by the local Drug Research Ethics Committees (18) and the Spanish Agency for Medicines and Health Products. All patients included signed written informed consent forms.

## Results

From February 1^st^, 2021 to September 30^th^, 2021, 311 patients were enrolled coinciding with the third to the fifth pandemic wave in Spain. Eleven patients withdrew informed consent prior to initiating treatment, resulting in a total of 300 patients in the analysis. Among these patients, the intention-to-treat, per-protocol and safety populations were equally constituted, with no major protocol deviations detected and all treatment doses received. The study protocol included an interim analysis with 50% of the estimated sample size, at which point the Scientific Committee decided to discontinue the study in light of the results presented below.

Of the 300 patients, 106 (35.33%) were assigned to the prophylaxis group (A group); 91 patients (30.33%) were allocated to the intermediate dose group (B group), and 103 patients (34.33%) were randomized to the therapeutic dose group (C group) (Flowchart in Supplementary file figure S1).

Baseline characteristics were similar in the 3 groups, including D-dimer, IL6, CRP, and ferritin values (Table 1). The distribution of individuals with D-dimer <1,000 μg/L was as follows: 83% in the prophylaxis group, 72% in the intermediate dose group, and 79% in the therapeutic dose group. Treatment for COVID-19 with corticosteroids (89.3%), remdesivir (18.0%), or tocilizumab (14.3%) was comparable in all 3 groups. The percentage of COVID-19-vaccinated subjects was was 16%, 29% and 26% in the prophylaxis, intermediate and therapeutic dose groups of tinzaparin, respectively (P = 0.06).

**Table 1.** Basal characteristics of patients included in PROTHROMCOVID trial.

| **<u>N= 300</u>** | **<u>Prophylaxis</u>**<br>**<u>(4500 UI</u>**<br>**<u>tinzaparin)</u>**<br>**Group A**<br>**(N= 106)** | **<u>Intermediate</u>**<br>**<u>(100 UI/ kg</u>**<br>**<u>tinzaparin)</u>**<br>**Group B**<br>**(N =91)** | **<u>Therapeutic</u>**<br>**<u>(175 UI/ kg</u>**<br>**<u>tinzaparin)</u>**<br>**Group C**<br>**(N= 103)** | **<u>P value</u>** |
| --- | --- | --- | --- | --- |
| Age, Media (SD) (years) | 54.1 (15) | 56.5 (14) | 58.5 (14) | 0.16 |
| Weight, Median (Q1-Q3) (Kg) | 79.6 (73-87) | 78.5 (70-88) | 78.9 (70-88) | 0.83 |
| BMI Median (Q1-Q3) | 28.5 (25-31) | 28.6 (26-31) | 28.7 (25-32) | 0.85 |
| Men, N (%) | 63 (59.4%) | 57 (62.6%) | 62 (60.2%) | 0.89 |
| Women, N (%) | 43 (40.5%) | 34 (37.3%) | 41 (39.8%) |  |
| **Cardiovascular risk factors** |  |  |  |  |
| Hypertension, N (%) | 29(27.4%) | 34(37.4%) | 36(34.9%) | 0.28 |
| Diabetes mellitus, N (%) | 13 (12.3%) | 17 (18.7%) | 20 (19.4%) | 0.32 |
| Dyslipidemia, N (%) | 26 (24.5%) | 30 (33.0%) | 36 (34.9%) | 0.22 |
| Smoking, N (%) | 5 (4.7%) | 6 (6.6%) | 5 (4.8%) | 0.82 |
| **Comorbidities** |  |  |  |  |
| Coronary heart disease, N (%) | 4 (3.8%) | 3 (3.3%) | 3 (2.9%) | 1.00 |
| Chronic obstructive pulmonary disease, N (%) | 3 (2.8%) | 4 (4.4%) | 5 (4.8%) | 0.76 |
| Chronic renal dysfunction, N (%) | 1 (0.9%) | 2 (2.2%) | 3 (2.9%) | 0.6 |
| Prior stroke, N (%) | 3 (2.8%) | 1 (1.1%) | ---% | 0.22 |
| Prior thromboembolic events, N (%) | 1 (0.9%) | 1 (1.1%) | 2 (1.9%) | 0.84 |
| **Respiratory severity** |  |  |  |  |
| SatO<sub>2</sub>/ FiO<sub>2</sub>, Median (Q1-Q3) | 353 (217-452) | 346 (199-450) | 342 (215-477) | 0.55 |
| **Laboratory test** |  |  |  |  |
| Peak D-dimer, Median (Q1-Q3) (µg/dl) | 618 (375-1100) | 686 (404-1340) | 620 (363-1200) | 0.63 |
| Platelets, Median (Q1-Q3) (x 10<sup>3</sup>) | 344 (269-436) | 369 (299-439) | 320 (246-401) | 0.07 |
| IL6* (Q1-Q3), Median (Q1-Q3) (mg/dl) | 23.8 (7.8-50.1) | 29.4 (5.7-63.8) | 21.43 (7.4-43.9) | 0.72 |
| Creatinine, Median (Q1-Q3) (mg/dl) | 0.76 (0.6-0.9) | 0.73 (0.6-0.8) | 0.71 (0.6-0.9) | 0.57 |
| Ferritin, Median (Q1-Q3) (ng/ dl) | 619 (274-1275) | 775 (386-1347) | 554 (271-1177) | 0.20 |
| CRP†, Median (mg/dl) | 57.6 (25-107) | 60.9 (14-142) | 57.1 (27-131) | 0.80 |
| LDH, Median (Q1-Q3) (ng/ dl) | 336 (254-439) | 333 (250-478) | 301 (243-383) | 0.049 |
| ISTH-DIC score‡, Mean SD) | 2.42 (0.9) | 2.56 (0.91) | 2.33 (0.76) | 0.19 |
| **COVID-19 Treatment** |  |  |  |  |
| Steroids, N (%) | 94 (88.6%) | 83 (91.2%) | 91 (88.3%) | 0.78 |
| Remdesivir, N (%) | 20 (18.8%) | 16 (17.5%) | 18 (17.4%) | 0.95 |
| Tocilizumab, N (%) | 16 (15.09%) | 18 (17.4%) | 11 (10.6%) | 0.35 |

Primary endpoint: The composite endpoint, ensued in 58 participants (19.3%) of the total study population; 19 patients (17.1 %) in group A, 20 (22.0%) in group B, and 19 (18.45%) in group C (p= 0.72). (Table 2).

**Table 2.** Primary and secondary outcomes among treatment groups in PROTHROMCOVID trial.

| **Primary Outcome** | **Standard prophylaxis**<br>**Tinzaparin**<br>**4500 IU/kg** | **Intermediate dose**<br>**Tinzaparin**<br>**100 IU/kg/day** | **Therapeutic dose**<br>**Tinzaparin**<br>**175UI/kg** | **P value** |
| --- | --- | --- | --- | --- |
| Primary endpoint (day + 30), N (%) | 19 (17.9) | 20 (22.0) | 19 (18.4) | 0.769 |
| **Secondary outcomes** |  |  |  |  |
| Death<br>N (%) | 2 (1.89) | 3 (3.30) | 3 (1.94) | 0.798 |
| Thrombotic event,<br>N (%) | 4 (3.77) | 2 (2.20) | 2 (1.94) | 0.742 |
| ICU admission,<br>N (%) | 7 (6.60) | 6 (6.59) | 10 (9.71) | 0.630 |
| High flow nasal cannula,<br>N (%) | 13 (12.26) | 14 (15.38) | 13 (12.62) | 0.786 |
| Noninvasive mechanical ventilation,<br>N (%) | 4 (3.77) | 4 (4.40) | 2 (1.94) | 0.668 |
| Invasive ventilation,<br>N (%) | 1 (0.95) | 2 (2.20) | 3 (2.91) | 0.602 |
| Progression WHO* scale,<br>Mean (Q1; Q3) | -0.43 (-1; 0) | 0.13 (-0.5; 1) | 0.06 (0; 1) | 0.689 |
| Progression to adult respiratory distress syndrome, N (%) | 4 (3.77) | 2 (2.20) | 1 (0.97) | 0.403 |
| Length of hospital stay,<br>Mean (Q1; Q3) | 16.4 (6.0; 17.0) | 15.0 (6.0; 24.0) | 13.9 (6.0; 14.0) | 0.962 |
| Major bleeding | - | - | - | - |
| Clinically relevant non-major bleeding, N (%) | 4 (3.77) | 3 (3.30) | 3 (2.91) | 1.000 |

The survival analysis revealed no statistically significant intergroup differences at 30 days. In all three groups, most survived to medical discharge without the appearance of the primary endpoint. (group A, 95% CI: 0.82 (0.73-0.88); group B, 95% CI: 0.78 (0.68-0.85); group C: 95% CI: 0.81 (0.73-0.88); Log-rank test P-value=0.75) (Figure 1). No differences were observed in survival when the groups were stratified according to D-dimer values (p=0.40)(Supplementary file Figure S2).

**Figure 1.**
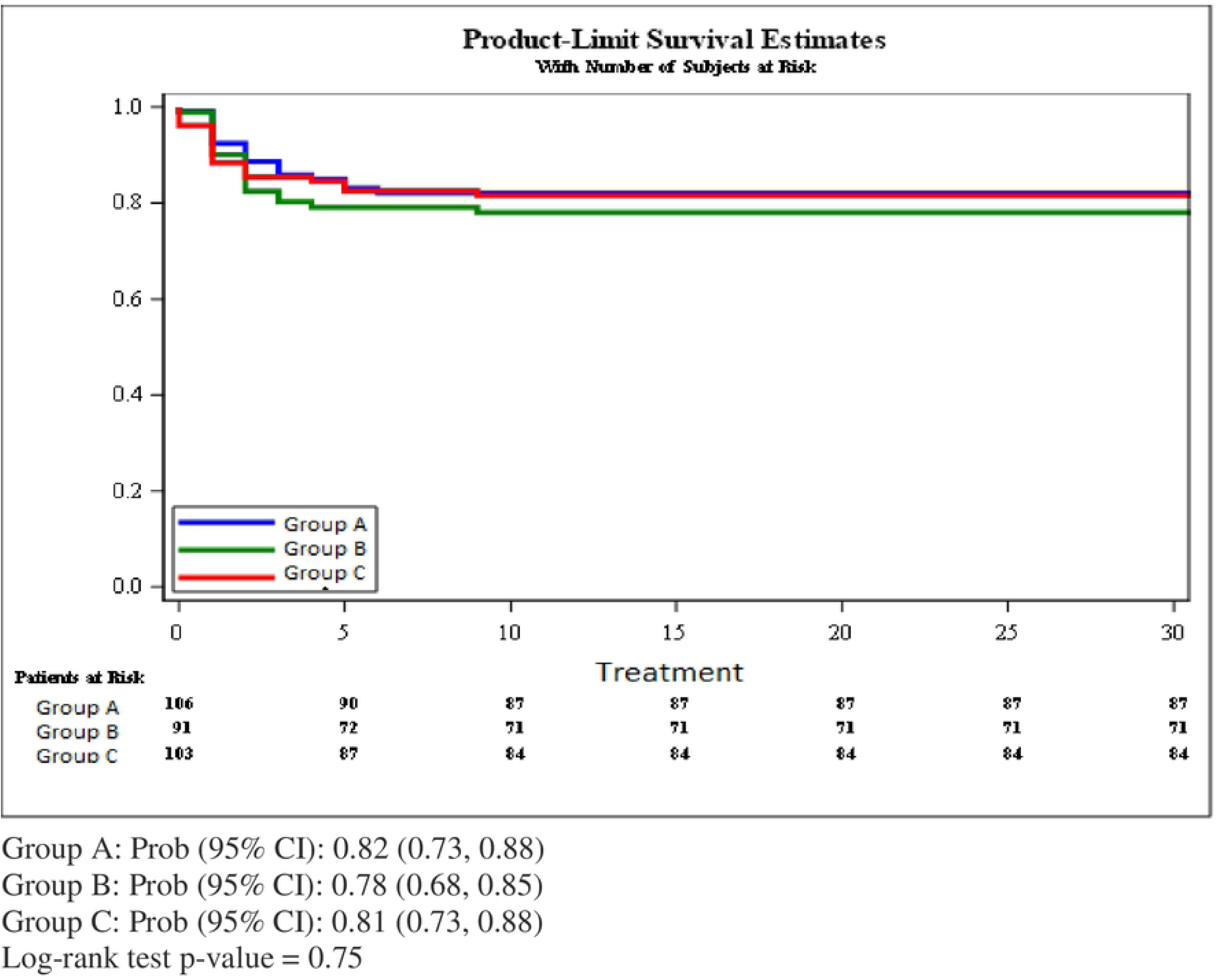
Overall survival of patient series, as per low molecular weight heparin group assignment. Group A, prophylactic dose (tinzaparin 4500 IU/ daily). Group B, intermediate dose (tinzaparin 100 IU/ kg/ day). Group C, therapeutic dose (tinzaparin 175 IU/ Kg/ day).

In terms of safety, the rate of bleeding was very low in all 3 groups. No major bleeding was reported and 7 patients (6.6%) in group A, 3 participants (3.2%) in group B, and 3 patients (2.9%) in group C suffered non-major bleeding, with no significant differences across groups (P=0.38). (Supplementary file Table S3)

Secondary endpoints: Secondary efficacy outcomes are displayed in Figure 2. A thrombotic event occurred in 4 patients in the prophylaxis group (3.8%); in 2 patients (2.2%) in the intermediate dose group, and in 2 subjects (1.9%) in the therapeutic dose group. NIV was provided for 10.5% of the participants in group A, 11.8 % in group B, and 4.9 % in group C. Seven (2.3%) of the included participants died during the first 30 days; 2 in the prophylactic dose group, 3 in the intermediate dose group, and 2 in the therapeutic dose group (P=0.48).

**Figure 2.**
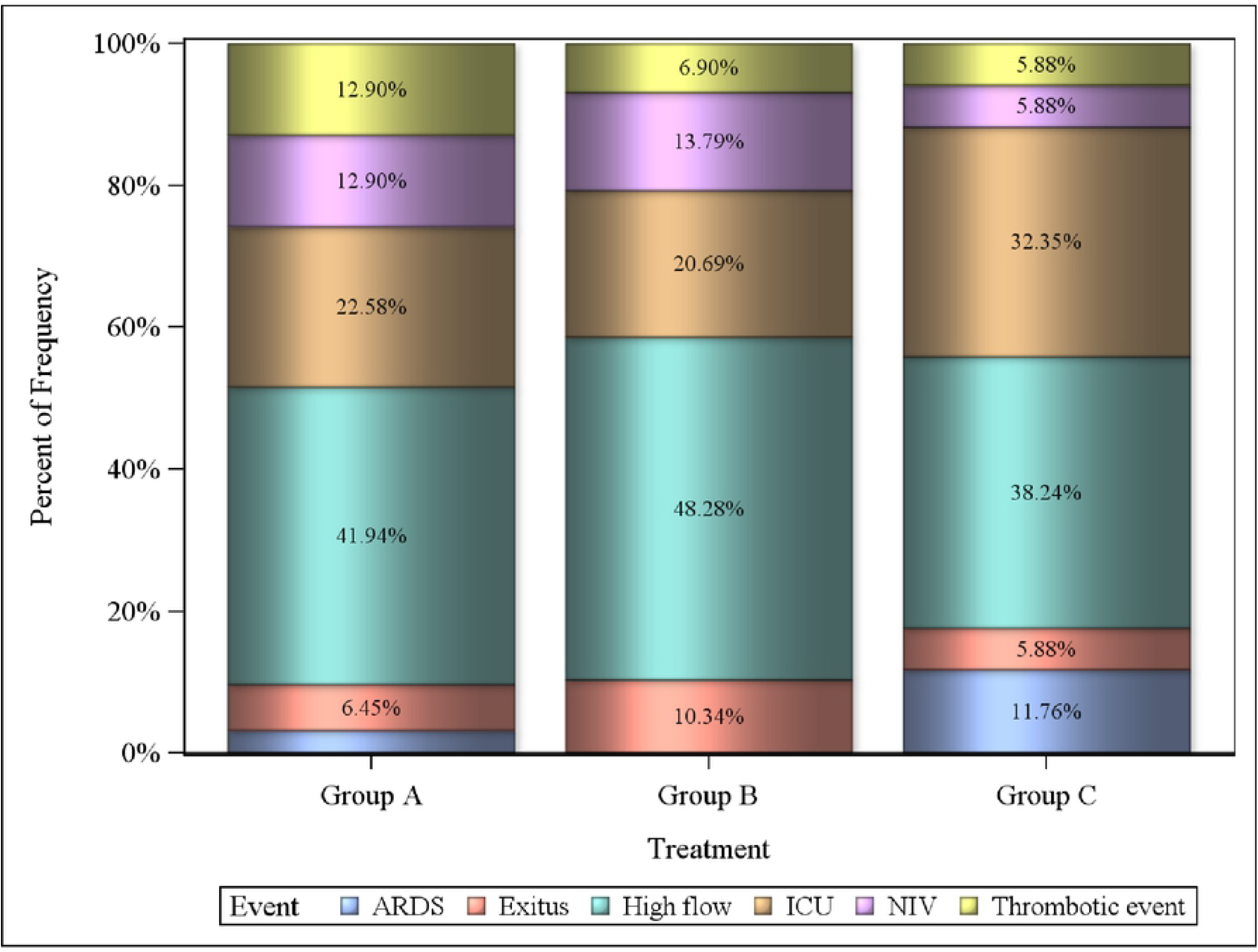
Frequency of the main secondary efficacy endpoints. *ARDS: Acute Respiratory Distress Syndrome. HFNC‡: High flow therapy with nasal cannula. ICU‡: Intensive Care Unit admission. §NIV: Non-invasive ventilation.

The World Health Organization (WHO) progression scale indicated no intergroup differences in progression between the date of admission and day 4, day 7, and at discharge. As for respiratory interventions, at day 4, 10% of the patients did not require oxygen therapy; 87% required oxygen therapy with nasal goggles or non-rebreather facemask; 0.74% required HFNC or NIV; 0.4 % needed NIV, and 0.4 % required IMV. The Wilkoxon paired signs test showed no differences in progression between groups.

Futility analysis showed that there were no evidences of significant differences between Groups A and B (Z=-0.71, P=0.48) and between Groups A and C (Z=-0.09, P=0.92); boundary values: α=-2.72; β=-0.70 (Supplementary file).

## Discussion

The results of the PROTHROMCOVID trial did not show differences during treatment with tinzaparin in relation to prophylactic, intermediate, or therapeutic doses in relation to the probability of death, thrombotic event, or non-invasive ventilation or invasive mechanical ventilation in patients with COVID-19 pneumonia. In this regard, the results of our trial provide evidence about the use of LMWH in non-critical patients with pneumonia due to COVID-19. This study tested these three strategies of different LMWH doses that coexisted *de facto* in different hospitals in the absence of solid evidence of the most suitable dose and the high rate of thrombosis and respiratory failure recorded in the first wave of the pandemic.

The results of PROTHROMCOVID trial are in line with a previous study published by Lópes et al. The ACTION trial, carried out an the end of first and second waves of the pandemic, included 615 patients and used a hierarchical statistical analysis structure based on time to death and detected no benefit in survival or in duration of hospitalization in individuals treated with full-dose enoxaparin or rivaroxaban compared to those who received standard prophylactic LMWH doses (11).

Similarly to our results, the RAPID trial determined that there was no significant difference between therapeutic or prophylactic strategies in non-critically ill patients admitted for COVID-19 in the combined endpoint of death, mechanical ventilation, or ICU admission (13). Moreover, the BEMICOP clinical trial, a small study conducted with bemiparin, did not find any differences in the primary endpoint between cases randomized to therapeutic doses in comparison with prophylactic doses (14). In contrast, the results of REMAP-CAP, ACTIV-4a, and ATTAC multi-platform collaborative trials support an early strategy of full-dose anticoagulant doses of heparin in non-critically ill subjects by demonstrating an increased in organ-free support days (98.6% vs 95.0%, respectively) in comparison to standard doses of LMWH thromboprophylaxis (10). Despite being the clinical trial that has included the largest number of patients, statistical significance was barely reached and there were no statistically differences in other outcomes among groups, including thrombosis, survival to hospital discharge and bleeding. Besides, the percentage of patients who received intermediate doses in the prophylaxis group was high (26%), which may have biased the results (15). The HEPCOVID trial, with a dose design similar to PROTHROMCOVID trial, showed a decrease in events, thromboembolism and death in the therapeutic-dose LMWH in hospitalized but not in ICU patients, with no differences at the intermediate-dose (16). It should be noted that the HEPCOVID trial was carried out in May 2020, during the first wave, with a higher percentage of events than the one observed in our trial and in those conducted in later stages of the pandemic. It is worth mentioning that PROTHROMCOVID recruitment began in February 2021, in the middle of the third wave of the pandemic in Spain and up to and including the fifth wave. Consequently, patients were at lower risk for mortality, given the widespread use of corticosteroids and the beginning of vaccination against SARS-CoV-2 (unlike other studies), with the first dose of tinzaparin administered within the first 24 hours and with concomitant treatment, mainly corticosteroids, most of which were homogeneous across the patients included. This profile is more similar to current clinical presentations than those of the first wave of the pandemic.

In line with our results, two clinical trials have analyzed standard prophylactic *versus* intermediate-dose LMWH. The INSPIRATION trial (17) tested the effect of intermediate versus standard dose prophylactic anticoagulation on thrombotic events, extracorporeal membrane oxygenation treatment, or mortality among patients with COVID-19 admitted to ICU. Likewise, Perepu et al (18) published the results of their trial that examined standard prophylactic versus intermediate dose enoxaparin in adults with severe COVID-19; both trials did not find significant intergroup differences. The lack of efficacy of the intermediate or full doses compared to standard doses could be due to differences in the clinical situation of the subjects included after the first wave, in which patients displayed more inflammation and received fewer doses of corticosteroids, monoclonal antibodies, immunomodulators and antivirals that have demonstrated benefit in the evolution of the disease. In contrast, the severity of individuals affected by SARS-CoV-2 variants with lower mortality rates in the last months of recruitment may account for these data. Similarly, the incidence of thrombosis recorded during the first wave (19) in our own setting was higher than data collected during the second wave.

A meta-analysis including 49 studies concluded that prophylactic anticoagulation was recommended against intermediate-to-therapeutic anticoagulation, considering insignificant survival benefits but higher risk of bleeding when higher doses were used. (20). The PROTHROMCOVID study confirms the non-superiority of intermediate doses with respect to standard prophylactic LMWH doses; consequently, the accumulated evidence suggests that this strategy should be abandoned.

However, the recommendations of the different guidelines have not been unanimous either. The American Society of Hematology favored prophylactic intensity over intermediate or therapeutic intensity anticoagulation for patients with critical illness related to COVID-19 or acute illness without confirmed or suspected thromboembolic disease (21), while the National Institute for Health and Care Excellence (NICE) guidelines put forth the conditional recommendation to consider a therapeutic dose of LMWH for young people and adults with COVID-19 who need low-flow oxygen and who do not have an increased bleeding risk (22).

In terms of safety, the risk of bleeding tends to be higher in most of the studies in which the anticoagulation strategy is more intense (23). In multiplatform trials, the risk of major bleeding was 1.8% in controls receiving standard prophylaxis versus 3.7% in those receiving therapeutic doses (10). The PROTHROMCOVID study participants had no major bleeding events, perhaps because of the smaller sample size than in the collaborative trials, the characteristics of the included population or the type of heparin used.(6)

We believe our safety data to be of the utmost importance because it does not appear from our results that the option of anticoagulation or intermediate doses generates an increased risk of unsafe major bleeding in the patient profile suggested by the NICE recommendation.

Our study has certain limitations; for instance, neither investigators nor patients were blinded. The main weakness of our results, however, is not having reached the estimated sample size of 600 patients, given that the researchers chose to interrupt the study on September 2021 because the results of the interim analysis. It revealed a very low absolute number of events in each arm and the futility analysis showed that it was unlikely that significant differences could have been reached with the complete sample. These results should not be extrapolated to other more severe hospitalized patients with COVID-19.

The strengths of our study include the low number of withdrawals of informed consent by patients, the very early use of tinzaparin in all three arms of the study, which may have influenced the favorable efficacy and safety outcomes and the fact that the three strategies of anticoagulation have been tested with the same LMWH. Similarly, this was a multicenter study conducted in academic and general care centers. Furthermore, the study was carried out during a phase of the pandemic were the incidence of thrombosis and mortality were lower than before; thus, the findings of our study might be more applicable to future waves of the pandemic, which are expected to be milder due to generalized immunization and better treatment options. (24).

In conclusion, in non-critically ill COVID-19 patients, intermediate or full-dose tinzaparin does not appear to offer any benefit over standard, prophylactic doses in the likelihood of thrombotic event, invasive or non-invasive ventilation, high flow oxygen with nasal cannula or death. However, administration of full or intermediate heparin doses in these patients is safe.

PROTHROMCOVID trial (NCT04730856).

## Data Availability

All relevant data are within the manuscript and its Supporting Information files. The data underlying the results presented in the study are available from https://prothromcovid.org/

www.prothromcovid.org

## Article Information

### Author Contributions

Dr. Muñoz-Rivas, Dr. Hernández-Rivas, and Dr. Torres-Macho had full access to all the study data and take responsibility for the integrity of said data and the accuracy of the data analyses. Both Dr. Muñoz-Rivas and Dr. Hernández-Rivas are corresponding authors.

*Concept and design:* Muñoz-Rivas, Hernández-Rivas, and Torres-Macho.

*Drafting of the manuscript:* Muñoz-Rivas, Hernández-Rivas, Méndez-Bailón, and Torres-Macho. The manuscript was critically reviewed by all the authors, who approved the final version.

*Statistical analysis:* Carlos Goetz and Juan Francisco Dorado, SAS Consultant from Pertica, SL.

### Other Information

The PROTHROMCOVID Investigators are listed in Suplementary material.

## Acknowledgments

We are indebted to the physicians, nurses, study coordinators, and data managers who contributed to PROTHROMCOVID clinical trial. We would also like to thank the patients and their families who participated in this study during the difficult times of the COVID-19 pandemic. The authors thank Leo Pharma for funding and support. The authors would also like to express a special thanks to Angélica Martin and Almudena Sánchez from S&H Medical.

